# Mechanisms for Integrating Real Data into Search Game Simulations: An Application to Winter Health Service Pressures and Preventative Policies

**DOI:** 10.1101/2023.09.14.23295499

**Authors:** Martin Chapman, Abigail G-Medhin, Kian Daneshi, Tom Bramwell, Stevo Durbaba, Vasa Curcin, Divya Parmar, Harriet Boulding, Laia Becares, Craig Morgan, Mariam Molokhia, Peter McBurney, Seeromanie Harding, Ingrid Wolfe, Mark Ashworth, Lucilla Poston

## Abstract

While modelling and simulation are powerful techniques for exploring complex phenomena, if they are not coupled with suitable real-world data any results obtained are likely to require extensive validation. We consider this problem in the context of search game modelling, and suggest that both demographic and behaviour data are used to configure certain model parameters. We show this integration in practice by using a combined dataset of over 150,000 individuals to configure a specific search game model that captures the environment, population, interventions and individual behaviours relating to winter health service pressures. The presence of this data enables us to more accurately explore the potential impact of service pressure interventions, which we do across 33,000 simulations using a computational version of the model. We find government advice to be the best-performing intervention in simulation, in respect of improved health, reduced health inequalities, and thus reduced pressure on health service utilisation.

## Introduction

Modelling complex real-world phenomena at an abstract level in order to gain initial insight into those phenomena is a common component of many scientific methods. As a part of this process, existing models—which can already capture features of the target phenomenon—are often used. This is particularly common in the field of game theory, where games like the *Prisoner’s Dilemma* have been used to conceptualise and understand potential solutions for problems in fields such as economics [1].

Another example of a game model that has been used in this way is a *search game*. In a search game, one or more *agents* (players in the game), *hiders*, conceal a set of objects in a discrete space (such as a graph), and a second group of one or more agents, *seekers*, must find these objects as efficiently as possible. A search game is suitably abstract so as to be able to represent a wide range of phenomena, but also suitably nuanced so as to be able to provide insight into specific circumstances. Like other game theoretic models, it is also amenable to computational realisation, allowing iterations of the game to be played out as, for example, an agent-based simulation or *digital mimic* (an artificial intelligence (AI) method), and the results recorded and analysed for insight. Search games (and their computational forms) have been used to investigate solutions to problems in areas like cyber security, where there are natural parallels between malicious actors on a network and both hiders and seekers [2].

Despite the nuance of a search game as a modelling tool, there is still often a disconnect between the abstract model and the real world, with significant validation needed before any potential solutions can be applied back to the original problem. To move towards bridging this gap, we propose the use of real data to configure search game models and thus make them more reflective of the phenomenon being modelled. We suggest four different types of data that should be used to configure a model: *demographic data*, which we segment into *individual* demographic data (providing input on agent state) and *distribution* demographic data (the spread of agent types); and *behaviour data*, which we segment into *primary* behaviour data (agent strategies) and *secondary* behaviour data (agent payoff).

To demonstrate the utility of our approach, we use a search game to explore solutions to the service pressures faced by healthcare providers during the winter months. Such a use case is timely given the current global cost-of-living crisis (COLC), which is exacerbating these pressures. We model a key driver of service pressures, which is respiratory and mental health conditions in children and young people (CYP) exacerbated by cold homes, and capture the government-backed (public health) policy interventions designed to address this. We then show how this abstract model can be configured using real data by drawing on six large datasets (including eLIXIR and Lambeth DataNet (LDN)) from the United Kingdom (UK), where these pressures are acute. From this model, we develop a simulation environment that is used to assess the impact of interventions in respect of improvements in health, a reduction in inequalities, and, ultimately, a reduction in pressure on health services such as the UK’s National Health Service (NHS).

## Background and Related Work

### Search games

Search games, in the simple form in which they were first introduced, define an abstract space ℛ into which a hider places an abstract *object* [3]. *We consider an evolution of this game and of the game of hide-and-seek* [4], *which focuses on the search for k* objects hidden on a network (*G* = (*V, E*), where *V* is a finite set of *nodes* (or *vertices*), and *E* a finite set of edges (paired vertices)). One agent, the seeker (*s*), is tasked with finding these objects which have been concealed by another agent, the hider (*h*). The task of the hider is to *plant* each object, and the set of nodes that is ultimately chosen by the hider as *hide locations* is defined as ℋ, where ℋ ⊆ *V*. In a multi-player variant of the game, hidden objects remain in place when discovered by a seeker, such that they can still be found by other seekers. The topology of the graph is arbitrary, and unknown by either player in advance. At the current point in a game, *t*, a player, *p*, is always associated with a vertex in *G*. When active, from their current vertex, a player is able to see 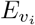; the set of edges leaving that node. By selecting one of these edges, they are able to move to adjacent nodes, and thus move through the graph in any direction (i.e. edges are bi-directional). The edges of this search space are *weighted*, so if a player moves along an edge it incurs the cost associated with its traversal.

The decisions made as to which node to move to next when at a given node is determined by a player’s *strategy*. As there are a number of different ways in which a player can make decisions, each player has a variety of different strategies available to them. Strategy names are prefixed by *s* and *h* to denote seeker and hider strategies respectively. Simple seeker strategies, such as a greedy strategy (*sGreedy*), might select the lowest-cost edge leaving a node, whereas more sophisticated strategies, such as a depth-first search (DFS), might approach the search space more methodically. More sophisticated strategies still might have a portfolio of strategies, Ψ, from which they initially select a strategy according to a given probability distribution, and then, based upon cues from the game about whether their choice is favourable or not, adapt or change strategy (subject to a confidence value, *λ*).

As a result of choosing a particular strategy, a player incurs a certain traversal cost. For a seeker, this reflects how efficiently they have located the hidden objects. It, therefore, makes sense to *reward* players in an amount that is inversely proportional to this value. Thus, the higher their traversal cost, the lower their reward. This reward is their *payoff* and is defined, for a seeker, in Definition 1, where *TCost* is a function mapping a player to the total cost of their graph walk. Under this payoff structure, players are motivated to visit each element of ℋ as efficiently as possible.

#### Definition 1

*A seeker’s payoff is inversely proportional to the total cost of their traversal: Payoff* (*s*) = −*TCost*(*s*).

### Winter pressures (WP)

It is well documented that during the winter months pressures on health services increase [5]. A significant contributor to these pressures is the poor health outcomes resulting from unsuitable living conditions. For example, homes often become cold and mouldy in the winter months if families cannot afford to heat them, and the children and young people (CYP) living there are susceptible to exacerbated chest conditions and mental health problems [6]. For families with a lower socioeconomic condition (SEC) these problems, and thus poorer health outcomes, are more pronounced. This is especially true during a cost-of-living crisis (COLC)—characterised by the prices of essential items, including food and energy, outpacing average household incomes—such as the global crisis that began in 2021 [7]. The United Kingdom (UK) has been significantly affected by the COLC, predominantly due to an inflation rate that reached a 41-year high in 2022 [8]. This has resulted in a marked increase in service pressures.

To offset these pressures, particularly during a COLC, governments often enact different types of preventative health policies. Key interventions include issuing advice (e.g. alternative methods to heat a home), support payments (e.g. household support funds [9]) and increasing vaccine eligibility (e.g. increasing age-based eligibility for child flu vaccinations under the UK’s NHS). However, the impact of these interventions is often variable [10].

### Methods

The challenge faced by a seeker in a search game is to collect a set of resources (hidden objects) in an efficient manner. The challenge faced by those families within a population during the winter months—in particular those where a CYP in the family has an existing condition that is likely to be exacerbated by a cold home and the effects that has—is often the same; a family must acquire, for example, the heat needed (a form of resource) for their home, and not exceed their financial means in doing so. In other words, families often face *resource problems*, a type of problem that search games are designed to explore. As such, it is natural to model families as seekers in a search game.

With this in mind, we examine the winter pressures (WP) use case in more detail to determine which other aspects are currently modelled by the game, or can be modelled by developing the game formalism further. We consider, specifically, families in the UK—both nationally and within specific regions—with CYP that have existing respiratory (e.g. asthma) and mental health (e.g. depression) conditions, and the impact of cold homes on these families. We consider families with different SECs (low, middle and high) and different ethnicities (White British and Minority Ethnic). We also look to model government interventions in respect of these conditions, and attitudes within a family towards these interventions. Our resulting model spans four key areas: the environment, the population, interventions and behaviour. This is elaborated upon in the following sections and summarised in Table 1.

**Table 1:** A summary of the elements of a search game used to model the winter pressures (WP) use case.

| Use case | Model |
| --- | --- |
| **Environment** |  |
| Non-winter | Random dispersal of resources |
| Winter | Dispersal of resources at maximum distance |
| **Population** |  |
| Families containing CYP with respiratory conditions | Game success impacts payoff ($\delta = 1$); random search |
| Families containing CYP with mental health condition | Game success impacts payoff ($\delta = 1$); random search with backtracking |
| Families containing CYP without condition | Games have less impact on payoff ($\delta < 1$) |
| Low, middle and high SEC families | Low, medium and high gas levels offset traversal costs |
| White British and Ethnic Minority families | Different gas levels offset traversal costs |
| **Interventions** |  |
| Government advice | Seekers adopt the strategy *sMaxDistanceFirst* |
| Support payments | Increase gas levels across seekers ($\beta_{PAY}$) |
| Increased vaccine eligibility | Decrease game success impact across seekers ($\beta_{VAC}$) |
| **Behaviour** |  |
| Uptake of targeted education | Meta-strategy $\{sRandom, sMaxDistanceFirst\}$ |
| Uptake of support payments | Distribution over $\beta_{PAY}$ |
| Uptake of vaccines | Distribution over $\beta_{VAC}$ |

## Modelling

### Environment

With families modelled as seekers, we set the distribution of resources in the game space to represent both winter and non-winter environments. To do this, we employ an abstract hider that uses two hiding strategies to reflect these environments: *hRandom* and *hMaxDistance*, respectively. Under the first strategy, resources are hidden at random, and under the second they are hidden with as many nodes between them as possible so as to intentionally maximise the path of the seeker. To observe the impact of these two hiding strategies first consider Figure 1a, which shows a game configuration where resources have been hidden at random, and as a result are (by chance) clustered around the seeker’s starting node (*v*_6_). The challenge here is, assuming the seeker moves anti-clockwise, relatively straightforward: only three nodes, with a total cost of 30, need to be explored in order to gather all resources. In comparison, in Figure 1b the *hMaxDistance* strategy has been employed and as a result the smallest number of nodes the seeker needs to explore is 6 (with a cost of 60) regardless of the direction they move in; a much more challenging search space. Therefore, we can view a less challenging search space (e.g. 1a), where resources are more straightforward to obtain, as a model of the non-winter months, and a challenging search space (e.g. 1b) as a model of the winter months, where it is harder to obtain resources. Returning to our example where heat is a resource this follows, as in the summer months heat—as (typically) available naturally from the climate—is more readily available, whereas in the winter months it is more challenging to obtain.

**Figure 1:**
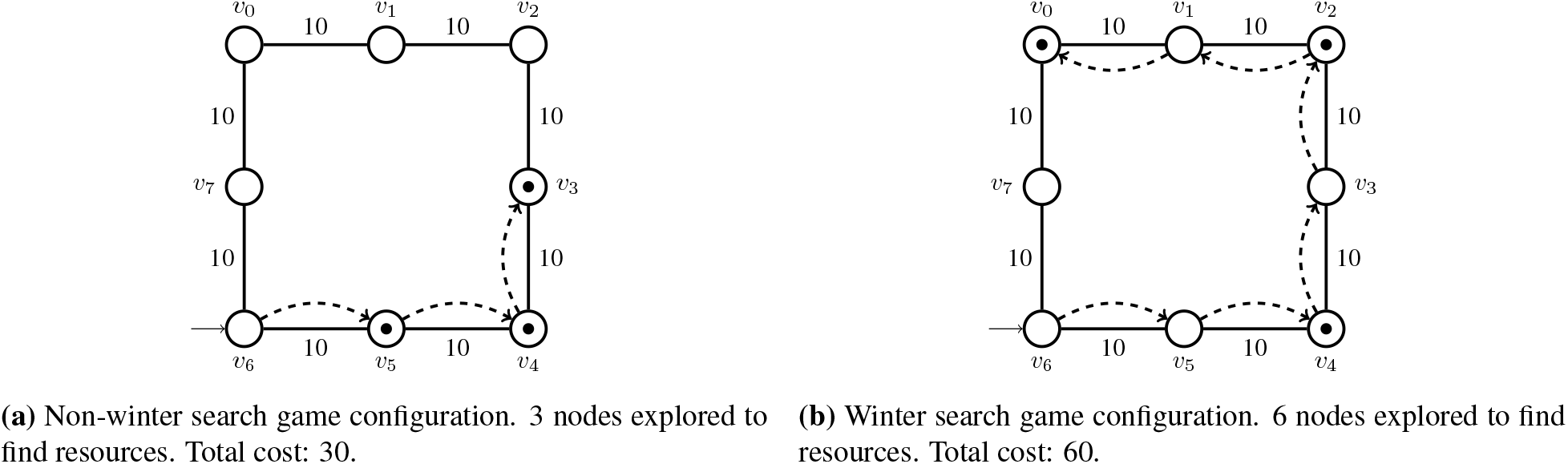
Representing non-winter and winter environments using a search game.

### Population

We next model the *state* of the families within a population. In the context of WP we consider three key state variables: (i) the presence of the aforementioned conditions in the CYP in that family (respiratory conditions and mental health conditions), (ii) a family’s SEC, and (iii) the primary ethnicity with which they identify.

We represent the presence of conditions in CYP by adjusting how payoff—defined previously as being inversely proportional to traversal costs—is derived in a game as follows:

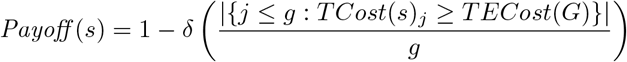

where *TECost* is a function mapping a graph to the total of its edge costs, and *g* is the total number of games. In other words, payoff now represents how ‘successful’ a seeker has been, on average, at collecting all their resources, and a seeker is successful in collecting all their resources if they reach all nodes containing a resource with a cost no greater than the total cost of all the edges in the space. Note also the constant *δ*, which we use to control the impact of ‘unsuccessful’ games on payoff (0≤ *δ*≤ 1). To demonstrate the idea of an unsuccessful game further, an example of one is shown in Figure 2a. Here, the seeker decides to backtrack upon having limited success during an initial search, and as a result the total cost of the search (100) is greater than the total sum of the edges in the graph (80).

**Figure 2:**
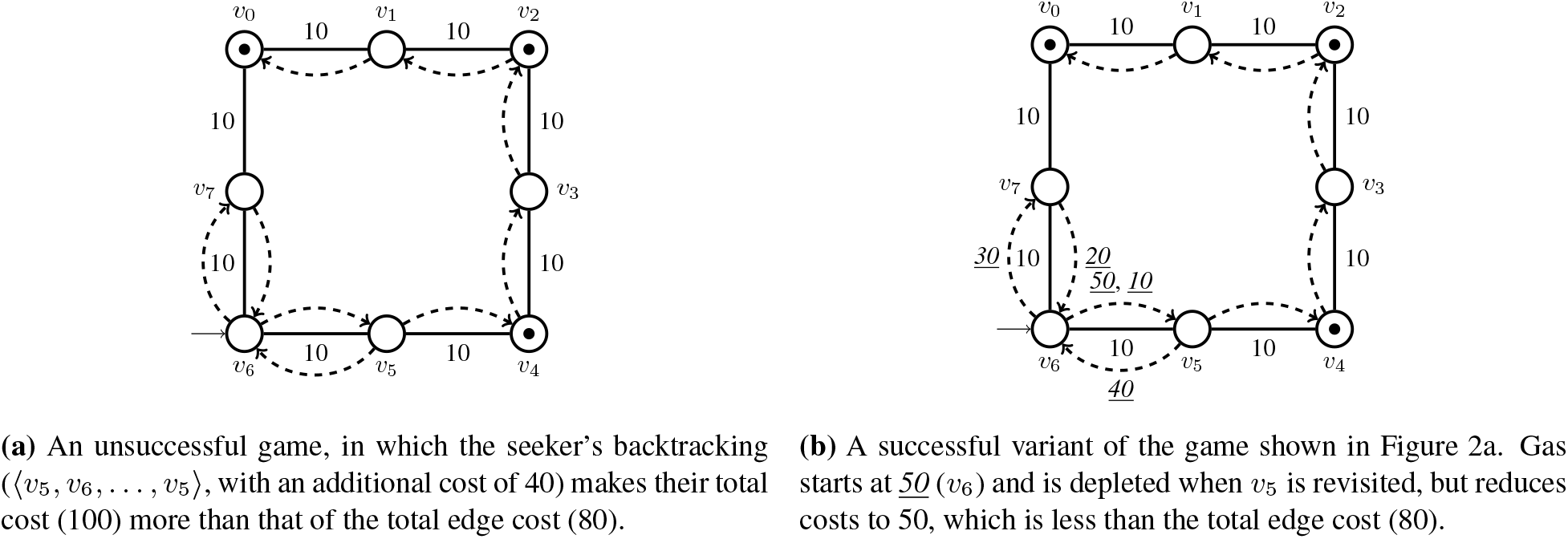
The impact of *gas* on an agent’s performance.

With this updated payoff mechanism in place the impact of not obtaining resources is better quantified, and if we set *δ* = 1 we are able to represent the fact that for families where children have conditions exacerbated by cold homes, failing to obtain the resources required (an unsuccessful game) has a direct impact on health outcomes (payoff). In the same way, setting *δ <* 1 allows us to represent, for example, families without CYP with any conditions, where the impact of a lack of resource acquisition is less marked. Indeed, in turn, because payoff now reflects health outcomes, it can also be used as a reasonable proxy for health utilisation, with a low payoff (poor health outcomes) suggesting higher health utilisation, and vice-versa.

To model different socio-economic conditions and different ethnic groups we introduce a concept into the search game model which we refer to as *gas*. Each player is given an amount of gas that is reduced when moving from node to node rather than a traversal cost being incurred directly. Once depleted, traversal costs are incurred as normal. The impact of gas on payoff is as follows, where *Gas* : *P* → ℝ^+^:

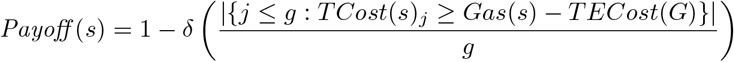

Figure 2b shows how the unsuccessful game shown in Figure 2a becomes a successful one with the introduction of gas to offset costs. Thus, in general, an agent with a higher initial amount of gas will be much more likely to have a successful game in a difficult resource environment. We therefore use varying gas levels to model families with different SECs, and because different ethnic groups are often characterised by a certain level of means ([11]), we also use gas levels to model different ethnicities.

### Interventions

We now consider how to model interventions using a search game. To represent each intervention (government advice, support payments and increasing vaccine eligibility) we leverage different aspects of the search game model discussed in the previous sections. When following government advice, an individual arguably updates an inherent ‘strategy’ that dictates how they approach a resource problem (e.g. utilising different mechanisms to heat a home). We reflect this in the search game model by updating the literal strategy used by an agent. To do this, we first need to select the default strategy used by agents in our model; those agents representing families with CYP with different conditions, and different SEC and ethnicity, as seen. As these groups are characterised predominantly by the updated payoff mechanism seen, the actual default strategy used to move from node to node is arbitrary. For all of these seekers, we therefore simply employ a random search strategy. The only exception is CYP with mental health conditions, where we employ a slight variant on the random search strategy which involves a level of backtracking, in order to reflect the impact these conditions may have on resource acquisition and to provide a means by which to differentiate these CYP from those with respiratory conditions. When enacting the government advice intervention within our model, we update this default strategy used by the seekers to be a targeted one designed to react to the *hMaxDistance* hiding approach used when representing a winter environment. This strategy is referred to as *sMaxDistanceFirst*, as it intentionally aims to visit nodes that are at maximum distance from one another, and thus avoid backtracking scenarios and additional cost. Because such a strategy aims to improve payoff by changing the way in which a resource problem is approached, it is reflective of the adoption of advice in the WP use case.

To model support payments and an increase in vaccine eligibility, we leverage the existence of gas and the *δ* constant introduced in the previous sections, respectively. In respect of support payments, we provide each agent with a baseline level of gas—in addition to the inherent level they have in order to represent SEC and ethnicity—in order to represent the additional means provided by such payments:

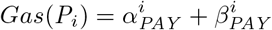

where 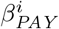 is the baseline gas of the *i*^*th*^ player, and 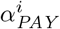 is the inherent gas level seen previously. We take exactly the same approach to modelling a vaccine eligibility intervention, where *δ* is now also derived as follows

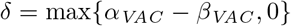

and the value of *β*_*VAC*_ can be increased to reflect the introduction of the intervention, as it reduces the impact that unsuccessful games have on payoff. This models how vaccines have the potential to align individuals to those without a condition, and reduce poor health outcomes.

### Behaviour

The final aspect of the WP use case to model is the behaviour of individuals in a population. We focus specifically on capturing behaviours in respect of the uptake of interventions. To model the uptake of government advice we introduce a meta-strategy that provides a distribution over the *sRandom* strategy (not using advice) and the *sMaxDistanceFirst* strategy (using advice). In other words, our *strategy portfolio* is

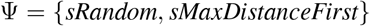

where the initial strategy chosen is determined by a given probability distribution, and we set *λ* (the confidence level, controlling further strategy change) to 0, to ensure only a single strategy is selected at the start of a game. We can, in a similar fashion, model variable uptake of support payments by introducing a probability distribution over 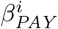 where 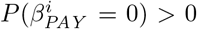, or, in other words, there is a chance that the additional baseline gas available will not be leveraged by a seeker. Finally, and once again in much the same way, we introduce a probability distribution for the values that comprise *δ* when deriving payoff, such that 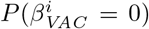, or there is a chance that the additional immunity available will not be leveraged.

### Data Integration and Results

In the previous section, we identified a set of parameters that can be used to configure an abstract search game model. In this section, we introduce four categories of real data that can be used to configure these parameters, with a view to increasing how closely a game models a target area. We present the results of analysing a variety of datasets in doing so, and use this data and the (evolved) search game model from the previous section to demonstrate the integration process in practice in the context of WP. All of the data resulting from the analysis conducted on these datasets is publicly available [12]. Following this, we demonstrate the specificity of the model by translating it to a computational platform and analysing the output of a series of simulations.

### Demographic data

#### Individual data

The first type of data that we deem important for configuring a search game model is data describing the demographics of the individuals being represented. This ensures each player’s state is set realistically, in accordance with the target domain. For example, in our WP search game model, an important aspect of a player’s state is their initial gas level, which is used to reflect families with differing SECs and ethnicities. Here, family income data can provide us with realistic gas levels. We sourced this data from the UK government (*n*=33,000) [13], and used high SEC and the ethnicity with the highest income in the dataset (in this instance White British) as a baseline to determine the other gas levels (for middle SEC, low SEC and Minority Ethnic) proportionally. This information is shown in Table 2. With this data integrated, we now have realistic gas levels that represent different SECs and ethnicities. Moreover, when combined with the different payoff mechanisms and strategies that model the presence or absence of certain conditions (described in the previous section), we now have distinguishable types of seeking agents in our model, e.g. one agent that represents a family with a low SEC and CYP with asthma (referred to using the naming convention *LowSEC-Resp*), and another that represents a family with a high SEC and CYP with asthma (*HighSEC-Resp*).

**Table 2:** Proportional income for different demographics.

|  | Proportion | Example gas |
| --- | --- | --- |
| SEC |  |  |
| High | 1 (baseline) | 500 (baseline) |
| Middle | 0.5 | 250 |
| Low | 0.22 | 110 |
| Ethnicity |  |  |
| White British | 1 (baseline) | 500 (baseline) |
| Minority Ethnic | 0.89 | 445 |

#### Distribution data

The presence of individual types of agents described in the previous section neatly prefaces the second element of data that is important for a search game which is distribution demographic data. This data is important to realistically set the spread of these different types of seekers in the model. In the WP search game model, this would be, for example, the portion of agents in the model that should be *LowSEC-Resp*, in accordance with the actual portion of these individuals found in the target population. To obtain these distributions, we combined data from three regional (South East (SE) London) studies: eLIXIR (Born in SE London), maternity and neonatal data from several NHS Trusts in London (*n*=32461) [14]; Lambeth DataNet (LDN), primary care data from 42 ethnically and socio-economically diverse GP practices in SE London, with an established data linkage with eLIXIR (*n*=66557) [15]; and Resilience, Ethnicity & AdolesCent Mental Health (REACH), mental health data for adolescents aged 11-18 years old in London (*n*=4000) [16]. To infer SEC in LDN we combined employment and Lower Layer Super Output Area information (LSOA; a fine-grained geographic segmentation mechanism for areas in the UK) with BMI and COVID-19 infection data, which are three measures that we deem to be a suitable proxy for explicit SEC data. In the remaining datasets, we extracted SEC and ethnicity data directly. Within LDN (and linked eLIXIR data) and REACH, we then broke down SEC and ethnicity further by our conditions of interest: respiratory conditions (asthma) and mental health conditions (depression), respectively. The resulting distributions are shown in Table 3.

**Table 3:** Derived distribution of demographics and conditions in our combined datasets.

|  | Regional |  |  | National |
| --- | --- | --- | --- | --- |
|  | Respiratory (*Resp*) | Mental Health | All conditions |  |
| SEC |  |  |  |  |
| High | 1.7% | 5.7% | 36.4% | 30.2% |
| Middle | 8.9% | 4.7% | 39.4% | 46.9% |
| Low | 4.9% | 1.0% | 24.1% | 22.9% |
| Ethnicity |  |  |  |  |
| White British | 4.5% | 2.1% | 23.0% |  |
| Minority Ethnic | 13.1% | 7.0% | 76.9% |  |

Translating these distributions to representative samples of 100 agents in our model, we constructed 15 groups of a size that reflects the actual size of these groups within the real population. Note that groups can be constructed based on different distribution types as well as the agent types seen previously. For example, in the WP use case we can also consider national studies, and their respective distributions, as well as regional studies in order to create distinct groups representing each population. To illustrate this, we combined national data on SEC again from the UK government (*n*=8589) with data from the Millenium Cohort Study (*n*=10757), a longitudinal UK dataset of CYP [17] (Table 3), and also used these distributions as input to our model.

### Behaviour data

#### Primary data

It naturally follows that in order to model the behaviour of individuals using a search game it is important to understand what those individuals would do in practice. As such, the third type of data we deem important for integration with a search game model is primary behaviour data. In the WP model, this data is important for modelling intervention uptake. That is, the distribution applied to the meta-strategy determining whether the *sRandom* or *sMaxDistanceFirst* strategies are chosen (used to reflect uptake of government advice) and the distributions for 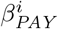 and 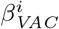 (access to payment and vaccine interventions, respectively).

To collect this data a survey questionnaire (primary data collection) was designed by the study team. The target group was identified as a broad cross-section of individuals living in the UK aged 18 years and above, with the goal of collecting views from the types of individuals (e.g. parents in a family) captured in our model. A draft of the questions was produced via discussion amongst a subset of the authors (A.G. and K.D.), with a subset of the remaining authors and external Patient and Public Involvement and Engagement (PPIE) advisors participating in an asynchronous iterative review process to critique and refine the questions into a final set. Participants were recruited using purposive sampling via dedicated PPIE platforms and links provided through funding and project partners. A copy of the questionnaire is publicly available (github.com/digitalmimic/winter-engagement). From the representative survey responses received (*n*=22), we were able to derive a set of distributions representing the likelihood of each intervention being accessed based upon positive and negative responses. Our distributions are 63%, 51%, 89% in favour of utilising government advice, accessing support payments and utilising vaccine interventions, respectively. With this data, we were then able to tune these distributions in our model, and ultimately ensure the behaviour of the agents is reflective of the individuals being modelled.

#### Secondary data

As there are often limitations to the scale of the data one can collect through primary data collection, the final type of data we identify as important for integration with a search game model is secondary behaviour data. In the WP model, this type of data is important to contextualise payoff. Recall that in our search game model payoff abstractly represents health utilisation (lower payoff models poorer health outcomes and higher utilisation). However, our payoff values themselves only make sense in the context of our abstract model. Therefore, real health utilisation (behaviour) data is needed to anchor these payoff values and provide meaning to any results. This data was extracted from LDN (primary care) and is shown in Table 4. Using these values, we can, by running a set of simulations of existing events (e.g. health utilisation in the non-winter months), establish a mapping between payoff and health utilisation, allowing us to derive accurate health utilisation values for predictive simulations.

**Table 4:**
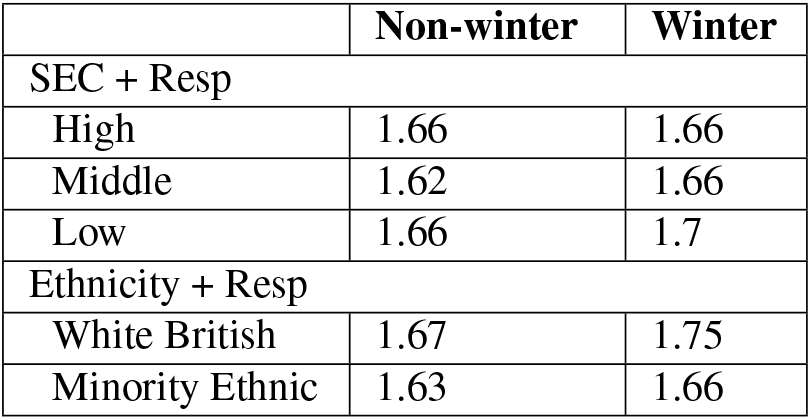
Primary care visits per year by demographic.

### Simulations

In the previous section, we outlined how real data was used to configure the state, distribution and behaviour of the artificial agents that comprise an abstract model in order to make it more representative of the WP use case. In order to further demonstrate the impact of using data in this way and the type of insight that can be gained by doing so, we can evaluate the payoff values produced by different groups of agents in our model in simulation. To do this, we first translated our model to a simulation platform using Java (github.com/digitalmimic/model). In total, using this platform, we ran 33,000 simulations across a variety of configurations; 1000 simulations for each configuration, a suitable point at which we expected to see a convergence of the results. The machine used for running the simulations detailed contained a 5.6 GHz 24-core processor and 64GB of DDR5 RAM. All pairwise values reported have been tested for statistical significance, with *p <* 0.01 in all cases unless otherwise stated.

As described in the previous section, we first ran a baseline set of simulations to construct a mapping between abstract payoff values and the real health utilisation data we have. We used a configuration representing health utilisation during the non-winter months for these simulations and focused on the *SEC-Resp* groups to demonstrate the process. Similar simulations could also be run for different demographics (ethnicity) and conditions (mental health). This baseline is shown in Figure 3 (striped bars). Leveraging this mapping, we then ran a simulation designed to reflect future winter months, the results of which are also shown in Figure 3 (transparent bars). Here, we can observe that the use of data allows us to accurately replicate several real-world trends of healthcare utilisation during the winter months. First, a key trend is the fact that there is an increase in utilisation across all groups—indicative of poorer health outcomes—which is less pronounced for the higher SEC group. Second, there is a disparity in health utilisation (health outcomes) between the groups, with the low SEC group being disproportionately affected. For completeness, and to ensure that these patterns are not due to extraneous factors, we removed data from our model by randomising all our parameters, and ran a third simulation, also shown in Figure 3 (solid bars). Here, the absence of the same real-world properties strongly suggests the accurate trends seen are introduced by the inclusion of data in our model.

**Figure 3:**
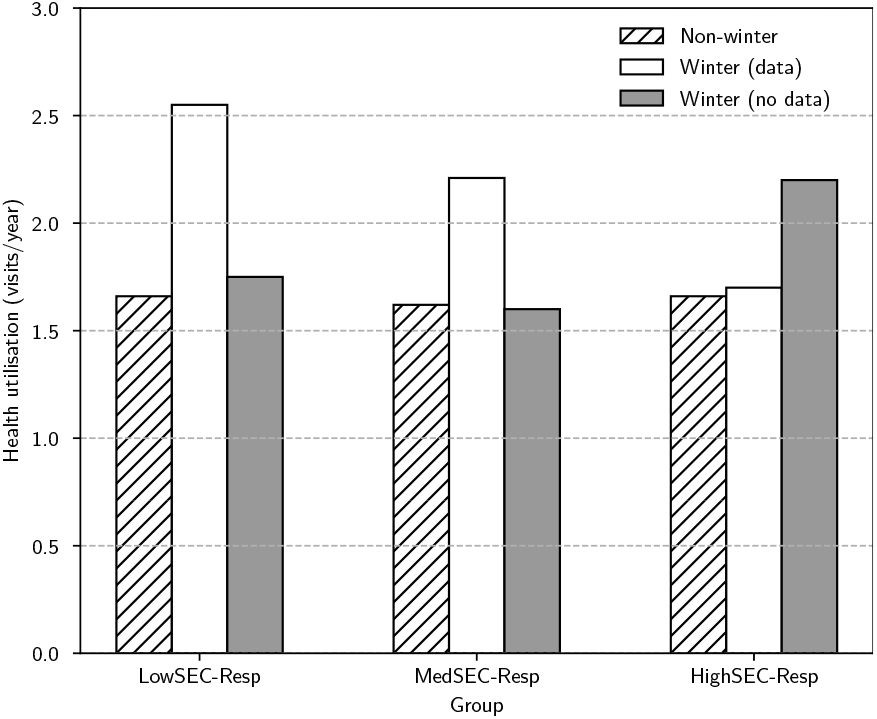
Health utilisation across different seasons and the impact of data.

Having shown the positive impact of data in allowing us to simulate the target domain accurately, we now present how government interventions can be simulated in our model and the type of insight gained. We again focus on the *SEC-Resp* groups as an example. We ran three winter configurations where the strategy used by the *SEC-Resp* agents is *sMaxDistanceFirst* (advice intervention), and set 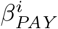 (payment intervention) and 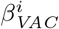 (vaccine intervention) to a moderate value (0.3), with a view to this acting as an independent variable in further simulations. The results of these simulations are shown in Figure 4. We can note several trends here which could be useful in policy decisions. For example, the overall best-performing intervention is government advice, both in respect of overall utilisation and reducing disparities between different SEC groups. In contrast, the remaining two interventions (payments and vaccinations) have limited impact and little difference is seen between them (0.01 *< p <* 0.05 for *LowSEC-Resp*).

**Figure 4:**
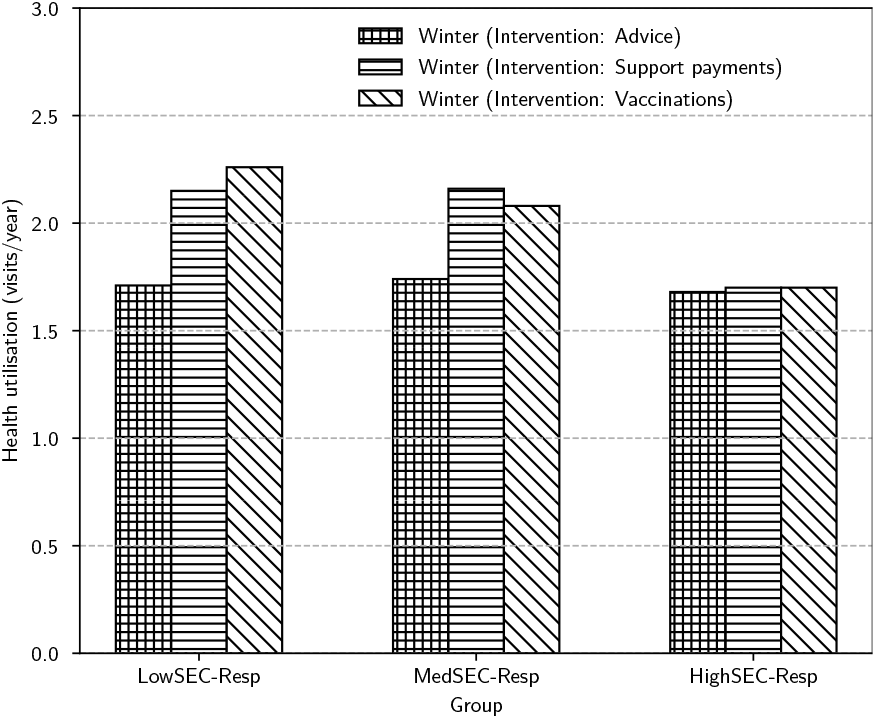
Projected impact of government interventions on health utilisation.

## Discussion and Conclusion

In this paper, we have shown the importance of combining search game models with real data when exploring complex issues in a domain. We have focused on a use case that centres around winter health service pressures and identified the types of data required to configure a model of this phenomenon, and others, accurately. To the best of our knowledge, this is the first application of search games to this use case. Using this configured model, we have shown the type of policy insight that can be gained by running it computationally. Specifically, we have shown the positive performance of government advice across different SEC groups, and the relatively poorer performance of the remaining interventions. There are a number of factors likely to contribute to this poor performance that we have captured in the model, including variable uptake of the interventions (something seen in the literature and our own data). With only one of the three interventions performing well, our overall interpretation of this result is that more fundamental systemic changes are needed that go beyond the interventions proposed in order to tackle winter health service pressures.

Despite the importance of using real data to configure aspects of a search game and the insight that can be gained by doing so, there are several caveats to consider. The first is that although the need for validation is reduced by the use of real data when configuring a model, this is not to say no validation is required at all. The results presented in the previous section, for example, would still require validation before implementation into policy. This could be done via a comparison with how interventions perform in practice once enacted. Secondly, the quality of a model is only as good as the integrated data. Without, for instance, suitably broad or complete data, the accuracy of a model may be negatively impacted. Finally, our emphasis on the importance of data is not to say that all parameters of a search game should be configured in this manner. A natural example of this is independent variables, such as the *β* variables in the previous section, where a variety of values are needed to explore payoff, and for which real data is not necessarily appropriate. In addition, there are often parameters in a search game which can be left as reasonable defaults. For example, when modelling WP, it was deemed suitable to leave parameters such as the number of hidden objects (10% of the total nodes) and edge weights (randomly weighted) as their default. Indeed, attempting to set too many variables could result in an unwieldy model. In general, which parameters need to be set by real data in a model in order to improve its accuracy (while leaving others as default to avoid unnecessary complexity) should be guided by key aspects of the domain being modelled.

Future work will look to build on the insight described above by integrating additional data into the model, such as additional SEC data in respect of intervention uptake (e.g. measured by group) and additional health utilisation data. Other future work will, in respect of the data integration process outlined, apply (i) the search game model to additional health domains, tailoring the model and sourcing relevant data under the same methodology that has been applied here; and (ii) examine other game theoretic formalisms for which real data is likely to be of use when modelling health domains, such as *flag coordination games*, which explore similar problems to that of the search game.

## Data Availability

All data produced are available online at

https://doi.org/10.5281/zenodo.8249580

## Acknowledgements and Ethical Considerations

This study is funded by the National Institute for Health and Care Research (NIHR). The views expressed are those of the author(s) and not necessarily those of the NIHR or the Department of Health and Social Care. This work also benefits from the infrastructure and partnerships assembled by HDR UK, including through the Data and Connectivity National Core Study, funded by UK Research and Innovation [grant ref MC PC 20058]. This research project was approved by the eLIXIR oversight committee in January 2023 (reference: DL038). Ethical approval for primary data collection was obtained through King’s College London (MRA-22/23-35470).

